# On the reliability of predictions on Covid-19 dynamics: a systematic and critical review of modelling techniques

**DOI:** 10.1101/2020.09.10.20192328

**Authors:** Janyce Gnanvi, Valère Kolawolé Salako, Brezesky Kotanmi, Romain Glèlè Kakaï

## Abstract

Since the beginning of the new coronavirus 2019-nCoV disease (Covid-19) in December 2019, there has been an exponential number of studies using diverse modelling techniques to assess the dynamics of transmission of the disease, predict its future course and determine the impact of different control measures. In this study, we conducted a global systematic literature review to summarize trends in the modelling techniques used for Covid-19 from January 1st 2020 to June 30th 2020. We further examined the reliability and correctness of predictions by comparing predicted and observed values for cumulative cases and deaths. From an initial 2170 peer-reviewed articles and preprints found with our defined keywords, 148 were fully analyzed. We found that most studies on the modelling of Covid-19 were from Asia (52.70%) and Europe (25%). Most of them used compartmental models (SIR and SEIR) (57%) and statistical models (growth models and time series) (28%) while few used artificial intelligence (5%) and Bayesian approach (3%). For cumulative cases, the ratio predicted/observed values and the ratio of the amplitude of confidence interval (CI) or credibility interval (CrI) of predictions and the central value were on average larger than 1 (4.49 ± 9.98 and 1.10 ± 1.94, respectively) indicating cases of incorrect predictions, large uncertainty on predictions, and large variation across studies. There was no clear difference among models used for these two ratios. However, the ratio predicted/observed values was relatively smaller for SIR models than for SEIR models, indicating that more complex models might not be more accurate for predictions. We further found that values of both ratios decreased with the number of days covered by studies, indicating that the wider the time covered by the data, the higher the correctness and accuracy of predictions. In 21.62% of studies, observed values fall within the CI or CrI of the cumulative cases predicted by studies. Only six of the 148 selected studies (4.05%) predicted the number of deaths. For 33.3% of these predictions, the ratio of predicted to actual number of deaths was close to 1. We also found that the Bayesian model made predictions closer to reality than the compartmental and the statistical models, although these differences are only suggestive due to the small size of the data. Our findings suggest that while predictions made by the different models are useful to understand the pandemic course and guide policy-making, there should be cautious in their usage.

## Introduction

The current outbreak of the novel coronavirus SARS-CoV-2, epi-centred in Hubei Province, China, has spread to many other countries [1] causing devastating public health impacts across the world. SARS-CoV-2 apparently succeeded in making its transition from animals to humans on the Huanan seafood market in Wuhan, China. Since March 2020, while new cases in China appears to have settled down, the number of cases are exponentially growing in the rest of the world [2] with Africa the least affected continent.

As with the two other coronaviruses that caused major outbreaks in humans in recent years (namely, Severe Acute Respiratory Syndrome and the Middle Eastern Respiratory Syndrome [3, 4]), Covid-19 is transmitted from human-to-human through direct contact with contaminated objects or surfaces and through inhalation of respiratory droplets from both symptomatic and a-symptomatic infectious humans [5].

In the absence of a safe and effective vaccine or antivirals, strategies for controlling and mitigating the burden of the pandemic are focused on Non-Pharmaceutical Interventions (NPI), such as social-distancing, contact-tracing, quarantine, isolation, and the use of face-masks in public [6]. Though many countries rely on those mitigation measures which help to slow the spread of the pandemic [7], scientists are actively engaged in efforts to understand the epidemiology of the disease. Modelling novel coronavirus disease has then become of extreme importance. Many researchers around the world have studied the patterns of Covid-19 pandemic and several mathematical, computational, clinical and examination studies have been put forward for modelling, prediction, treatment and control of the disease [6, 7, 8, 9, 10, 11, 12, 13, 14, 15, 16]. This growing interest of scientists has resulted in a deluge of studies predicting the dynamics of Covid-19, and summarizing trends in these studies is necessary. Some studies (e.g. [8, 17]) reported the difficulty of current models to accurately predict the Covid-19 pandemic. [17] showed that non-identifiability in model calibration using data on confirmed cases is a main source of large variation in model predictions. Other studies [8, 18, 19] have raised the issues of data quality that is necessary for accurate predictions. The type of models used could also affect the accuracy of predictions [11, 20].

Here we conducted a systematic and critical review of studies published between January 1st and June 30th 2020 on Covid-19 to (1) summarize trends in the modelling techniques used to predict Covid-19 cases and deaths, and (2) assess the reliability of predictions of Covid-19 cases and deaths. The overarching goal is to determine and discuss to what extent studies accurately predict Covid-19 cases and deaths and whether some differences exist among modelling techniques.

## Methods

### Article search and selection

PubMed, medRxiv and Google Scholar were accessed to search for studies. The following keywords were used: “Coronavirus”, “Covid-19”, “Corona”, “SARS viruses” and “2019-nCoV” separately and in combination with “Model/modelling”, “Prediction/Predicting”,”Dynamics” and “Forecast/Forecasting”. The time period covered was from January 1st 2020 to June 30th 2020. The bibliographies of retrieved studies as well as bibliographies of current reviews and texts were searched for additional relevant studies. The studies included are those related to Covid-19 and are model-based. All studies that did not include a model were excluded as were non-English language studies. We followed the PRISMA flowchart (see Figure 1) for studies screening and selection for the review. From an initial list of 2170 studies, 148 were finally included in the review (Figure 1).

**Figure 1:**
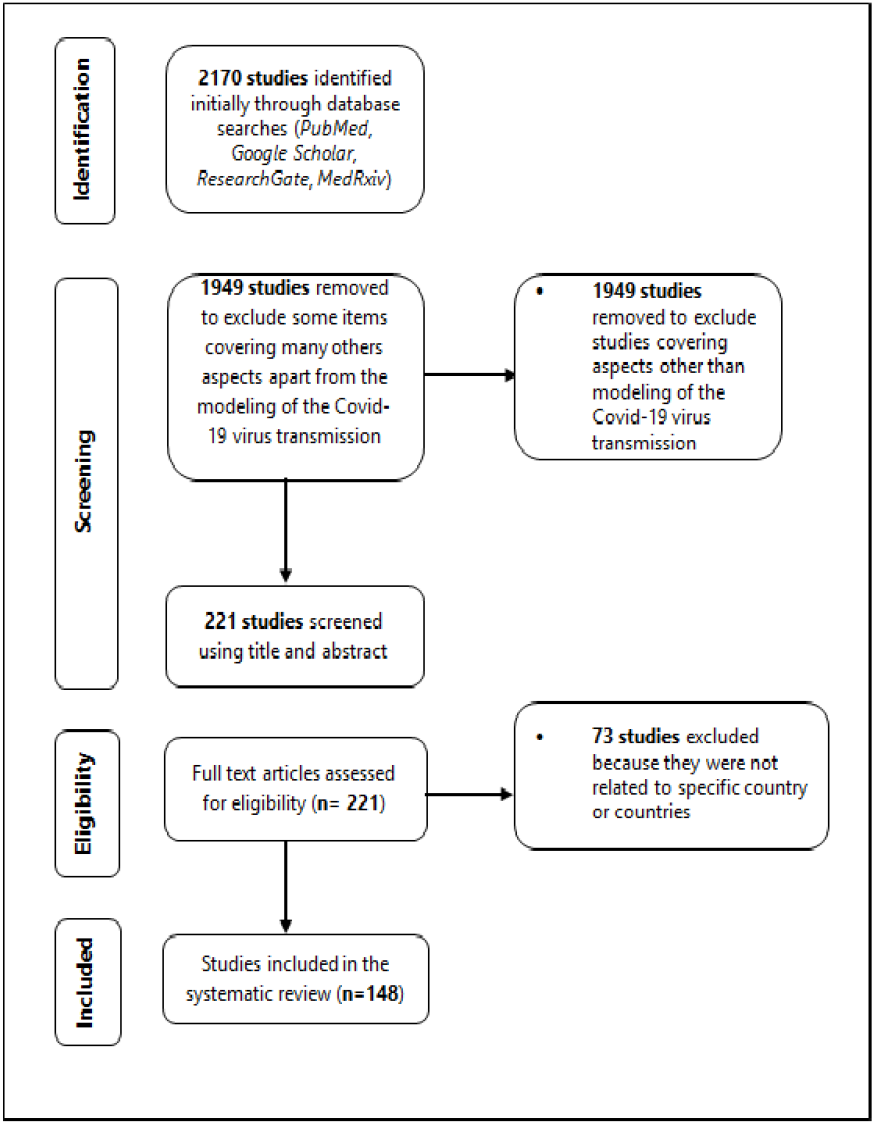
PRISMA flow diagram of the selection process for inclusion of studies in the review

### Literature synthesis and analysis

For each of the 148 papers selected, the data extracted were: the country of the study, the published or unpublished status of the study, the time period covered by the data (in days), the topics addressed in the study, the modelling techniques used, the predicted values of the cumulative number of cases, the predicted values of the cumulative number of deaths, the date at which the predicted values will be observed, and the uncertainty parameters (95% CI or 95% CrI). The data analyses considered three aspects. The first aspect was related to publication status of the study (published or preprint), geographical coverage (continents and number of countries covered per continent) and topics addressed in the studies; we used count and relative frequencies to describe this trend. The second aspect was related to the modelling techniques and was also addressed using count and relative frequencies after grouping modelling techniques in relatively similar groups. The third aspect was the reliability of the predictions made by studies. For this, we used four parameters; the first is the ratio between the value predicted and the actually observed value on the day on which the prediction was made. A value close to 1 indicates that the prediction was correct. Values less or larger than one indicate underestimation or overestimation, respectively. The second parameter is the ratio between the amplitude of the uncertainty parameter and the central value. For studies that used statistical methods, the uncertainty parameter is the 95% confidence interval (CI). For the studies which used Bayesian methods, the credibility interval is the uncertainty parameter and indicates that given the observed data, the effect has 95% probability of falling within this range. This ratio is an estimate of the accuracy of the predictions. A value of 1 for this ratio indicates that the amplitude is larger as the central value. Smaller values indicate accurate prediction (i.e. prediction with low uncertainty). The values of these ratios were plotted against studies, and type of models used. We additionally plotted these ratios against the number of days covered by the studies. We expect that the longer the period of data considered, the smaller the values of these ratios. The third parameter was whether the value actually observed in a study was within or out of the CI or CrI of the predictions; this was used to compute the proportion of studies for which observed data are within the CI or CrI of the predictions.

## Results

### Characteristics of the papers selected: publication status, geographical coverage and topics addressed

Of the 148 papers reviewed, 65.54% were preprints. The largest part of the studies were related to Asia (52.70%), especially to China (50) and India (14) (Figure 2 - (a)). However, the coverage (percentage of countries in a continent where a study was carried out) was higher in Europe where we found studies conducted in 77.3% (34 countries out of the 44) of European countries with Italy (25 studies), France (20 studies) and Spain (15 studies) being the countries where more researches have been done. From our sampled studies, only 17 focused on Africa either at country level (5 in Nigeria, 4 in South Africa and 2 in both Algeria and Kenya) or region level (i.e. west, east, north, south, or central), or the whole continent level. Some studies did not focus on a specific country but on an entire continent or part of continent (e.g [7]). Our selected studies covered 7 out of 35 countries in the America continent, with the Unites States (15 studies) and Brazil (6 studies) as the countries where most of the studies were carried out (Figure 2 -(b), Appendix).

**Figure 2:**
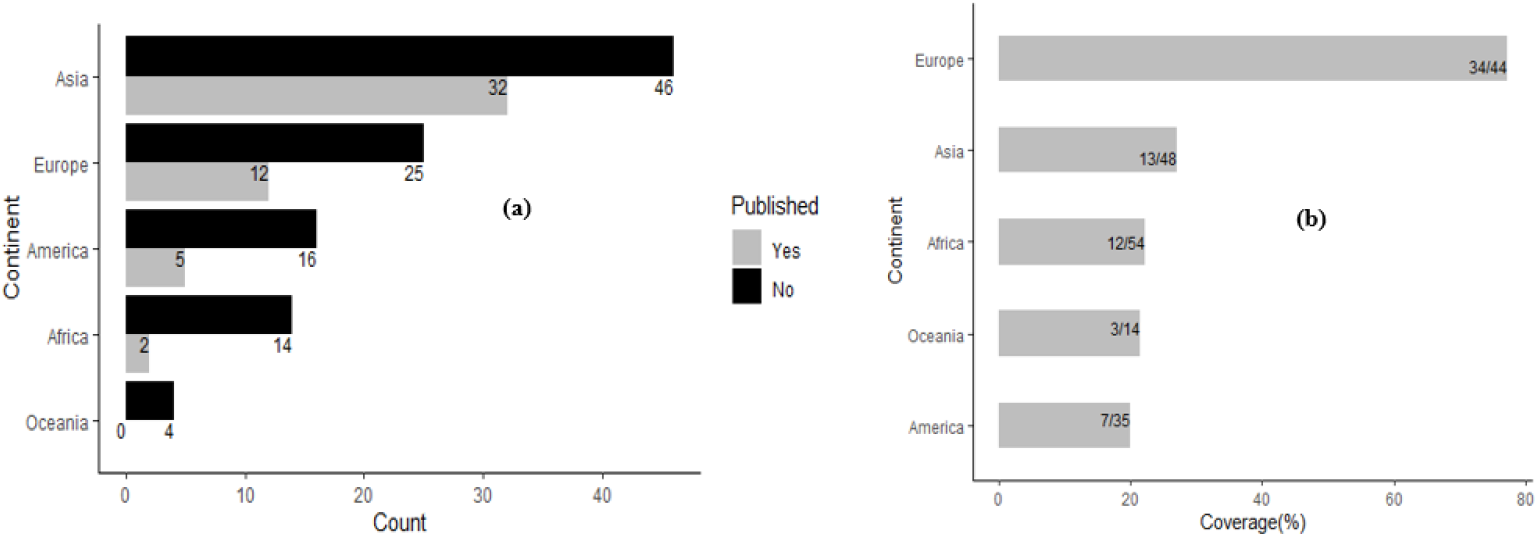
Countries coverage (in %) across continents

The studies addressed diverse topics which can be classified into four groups. In addition, the studies were primarily designed for three main purposes: study the dynamics of the transmission of Covid-19 with or without attempting to predict the course of the pandemic (cumulative cases, deaths, hospitalized cases etc.) (49.32% of studies); estimate key epidemiological parameters of the transmission of COVID-19 (57% of studies), and evaluate the impact of control measures on the transmission of Covid-19 (41% of studies). Other topics were covered (e.g. the burden of healthcare systems or healthcare resource estimation) but were classified in the category “Others”, due to their low proportion (6.76%) (Figure 3-(a)). With regards to the impact of control measures on the transmission of Covid-19, the following eight control measures were considered in the studies: face mask, quarantine, case isolation, contact tracing, social distancing, school closure, workplace distancing and restriction on international air travel. Figure 3-(b) shows the distribution of studies that assessed the impacts of the above control measures. The most assessed measures were quarantine (27%), social distancing (18.2%) and case isolation (12.2%). Eight epidemiological parameters were estimated (Figure 3-(c)) with the reproduction number, the most estimated parameter (41.22%).

**Figure 3:**
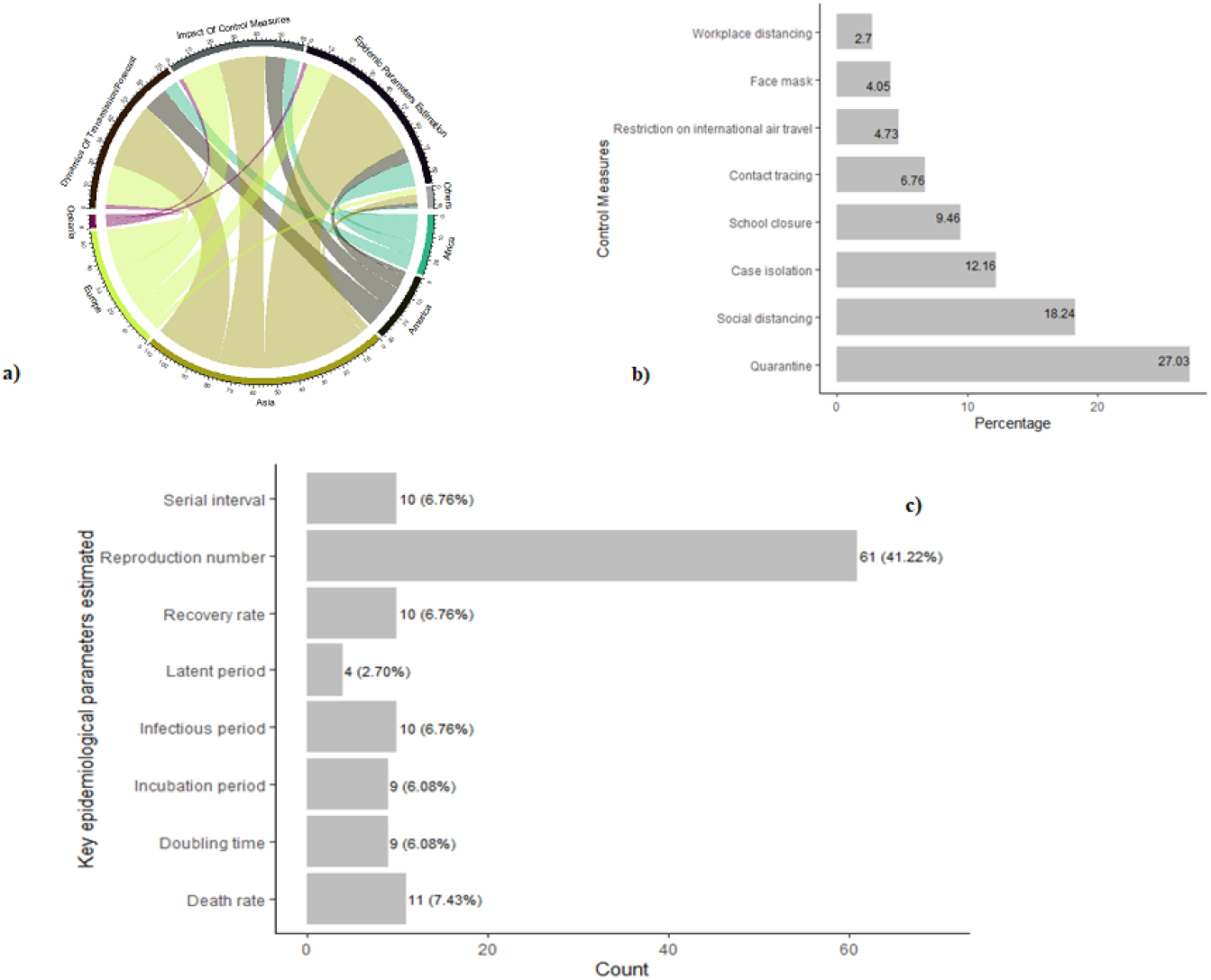
**a)** Plot of models used with the covered topics for the 148 studies **b)** Distribution of studies that assessed the impacts of the above control measures **c)** Distribution of studies that estimated key epidemiological parameters

### Modelling techniques

Several modelling techniques were used which we classified into five main groups, including Artificial Intelligence (AI) based approach, Bayesian models, Compartmental models, Hybrid models and Statistical models; they refer to models that combine two or more approaches. Hybrid models were considered by only 1% (2/148) of the studies and were a combination of a compartmental and statistical models and a combination of a compartmental and AI models. Compartmental and statistical models were the most used in the studies (57% and 28% respectively) (Figure 4). Compartmental models are extensively used regardless the topic addressed and are usually combined with statistical models (Figure 5).

**Figure 4:**
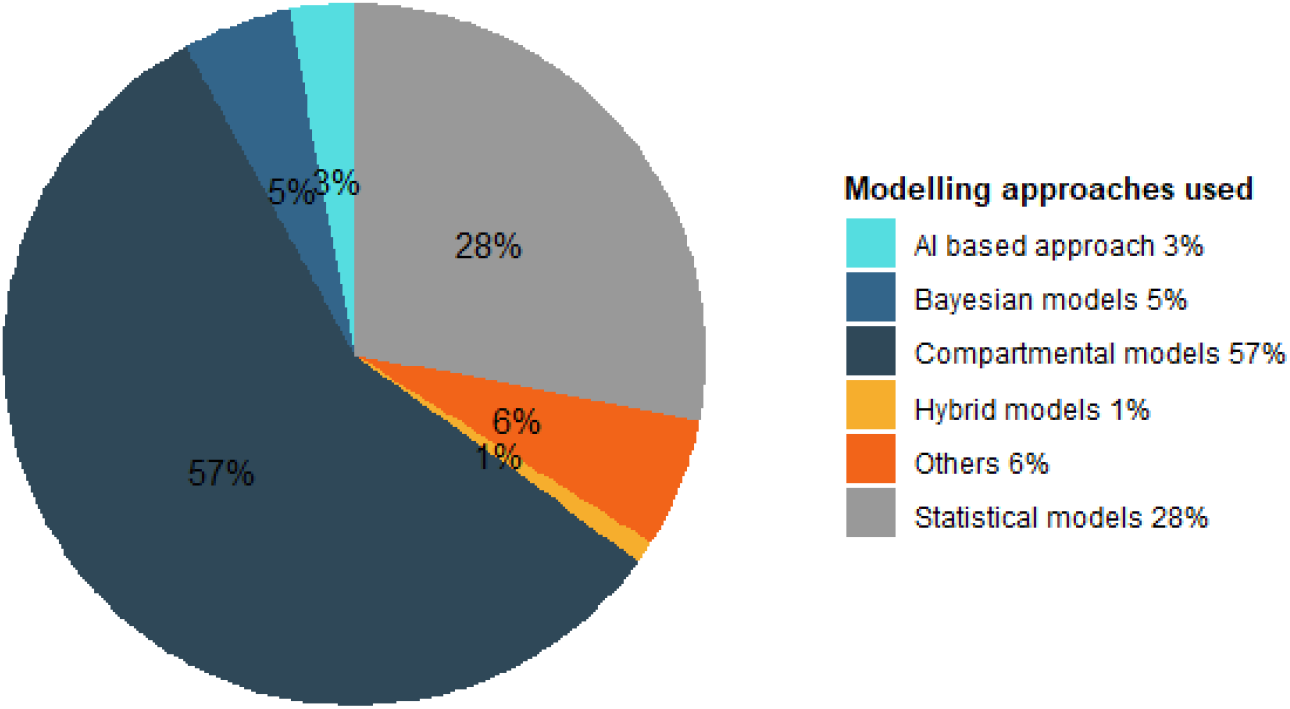
Diversity of modelling techniques used for Covid-19

**Figure 5:**
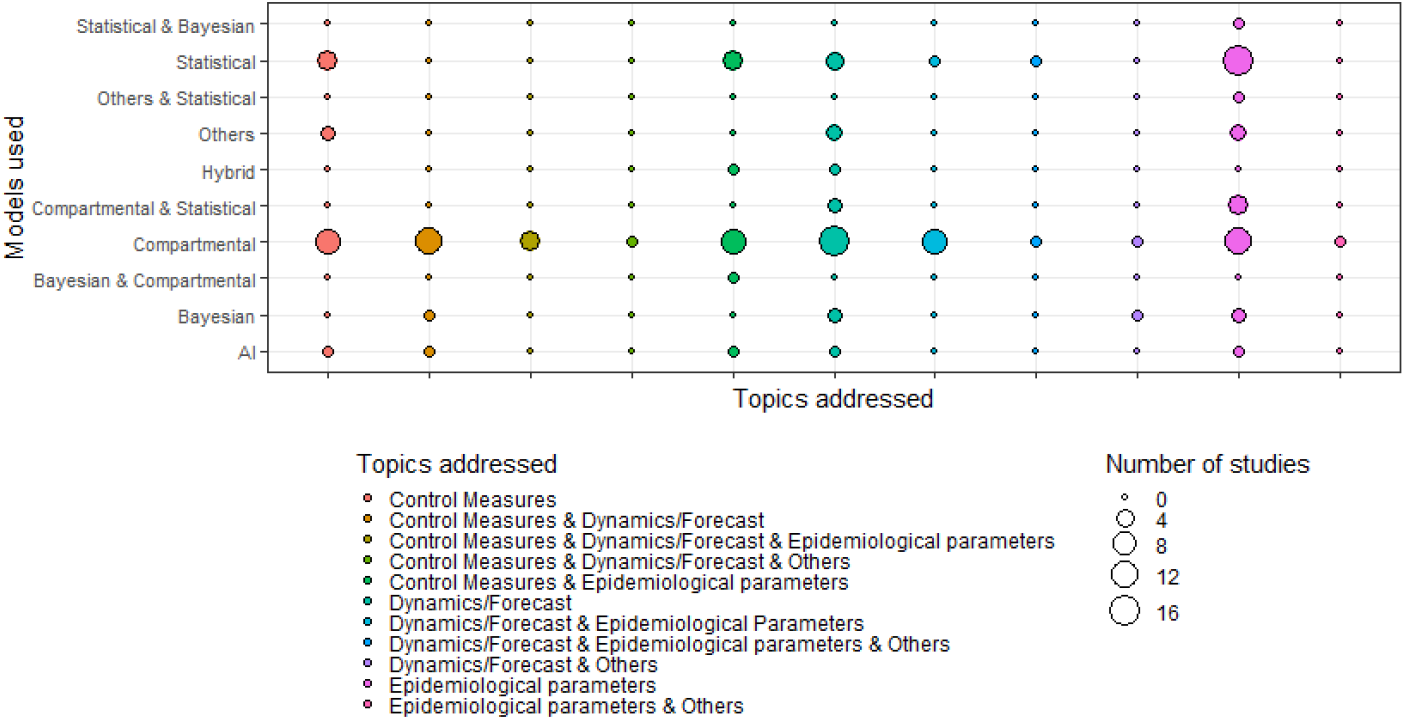
Topic addressed by modelling techniques of Covid-19

Overall, compartmental and statistical models were the most used models. A compartmental model is a model that stratifies the population into different compartments, such as different health states (e.g. Susceptible, Exposed, Infected, Quarantined, Recovered, Dead, etc.) for the modelling. Compartments are assumed to represent homogeneous sub-populations within which the entities being modeled–such as individuals or patients–have the same characteristics [21]. The classical SEIR model was considered in many studies (e.g. [22, 7, 23]) (22.30%). Some studies however adopted an improved SEIR model. Considering the traditional SEIR model as not realistic, [24] considered an improved SEIR model by introducing both quarantine status and intervention measures. [25] incorporate three important elements of Covid-19 to the classical SEIR model to estimate epidemiological parameters of the disease in South Korea, Italy, and Spain: (1) the number of asymptomatic infectious individuals (with very mild or no symptoms), (2) the number of symptomatic reported infectious individuals (with severe symptoms), and (3) the number of symptomatic unreported infectious individuals (with less severe symptoms). The SEDQIR model based on SEIR model was established by [10] with D, the suspected cases of infection or potential victims and Q, the diagnosed and quarantined. Moreover [26] introduced a compartment C and then developed the SEICR model.The compartment C was for confined individuals, that was, individuals whose movement are restricted and effectively removed from the susceptible population by strong NPI, like lock downs, closure of retail and entertainment, parks, and vehicular restrictions[26].

Statistical modelling is an approach for developing and testing theories by way of causal explanation, prediction, and description. In many disciplines there is near-exclusive use of statistical modelling for causal explanation and the assumption that models with high explanatory power are inherently of high predictive power [27]. Contrary to the compartmental models, statistical models often model one state (i.e. one compartment at the time) and do not often consider the flow of individuals among the different states. The most used statistical models were growth models which aimed to model change over time. About eighteen percent (17.6%) of studies used growth models (Exponential growth model, Generalized-growth model, Logistic growth model, Richard growth models etc.) [20, 28, 29]. About tenth of the studies used time series models (9.46%). Time series models that were used included ARIMA models [11], VAR models [30], exponential smoothing models [31] etc. One of the most common statistical modelling techniques are the regression models. Statistical models, such as regression models, are typically phenomenological and describe the statistical relationship or association between different model variables [21]. About six percent (5.4%; 8 out of 148) of the studies used a regression model (linear regression, polynomial regression etc.) [32, 33]. Additional modelling techniques were used, including parametric distributions fitting models [15, 34], exponential decay model [9], least square error (LSE) model [35].

**Table 1:**
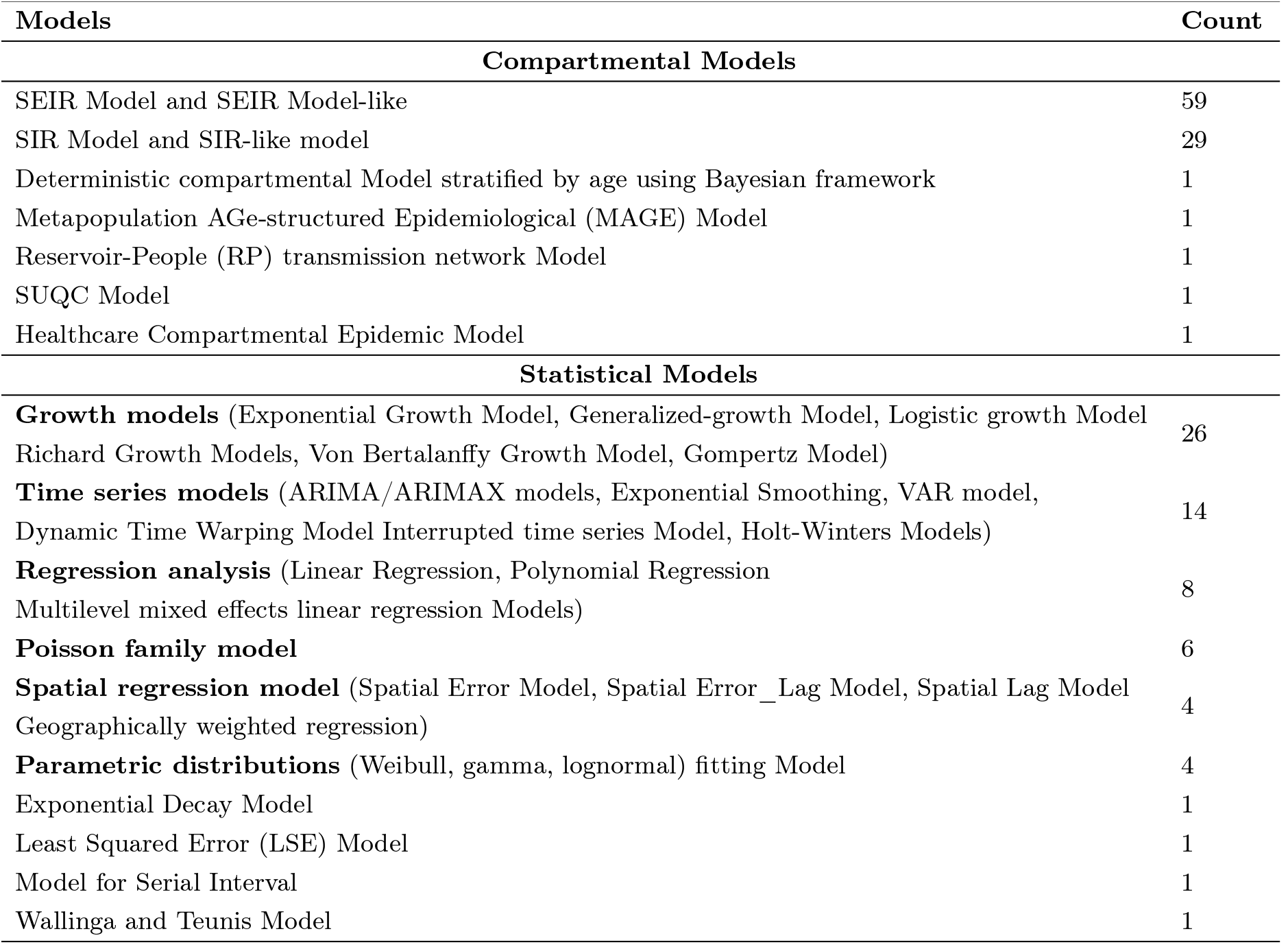
Compartmental and statistical models developed in the 148 studies

| Models | Count |
| --- | --- |
| **Compartmental Models** |  |
| SEIR Model and SEIR Model-like | 59 |
| SIR Model and SIR-like model | 29 |
| Deterministic compartmental Model stratified by age using Bayesian framework | 1 |
| Metapopulation AGE-structured Epidemiological (MAGE) Model | 1 |
| Reservoir-People (RP) transmission network Model | 1 |
| SUQC Model | 1 |
| Healthcare Compartmental Epidemic Model | 1 |
| **Statistical Models** |  |
| **Growth models** (Exponential Growth Model, Generalized-growth Model, Logistic growth Model, Richard Growth Models, Von Bertalanffy Growth Model, Gompertz Model) | 26 |
| **Time series models** (ARIMA/ARIMAX models, Exponential Smoothing, VAR model, Dynamic Time Warping Model Interrupted time series Model, Holt-Winters Models) | 14 |
| **Regression analysis** (Linear Regression, Polynomial Regression, Multilevel mixed effects linear regression Models) | 8 |
| **Poisson family model** | 6 |
| **Spatial regression model** (Spatial Error Model, Spatial Error_Lag Model, Spatial Lag Model, Geographically weighted regression) | 4 |
| **Parametric distributions** (Weibull, gamma, lognormal) fitting Model | 4 |
| Exponential Decay Model | 1 |
| Least Squared Error (LSE) Model | 1 |
| Model for Serial Interval | 1 |
| Wallinga and Teunis Model | 1 |

The dynamics of Covid-19 transmission have been analyzed in numerous studies which have tried to predict the spread of the disease using the above-mentioned models. While the predictions made by some of the models used are close to the observed reality, predictions made by other models have proven to be inaccurate. For example, on the basis of data from February 25 to March 21 and using the “Alg-Covid-19” Model, [36] estimated that the number of cases in Algeria will exceed 1,000 on the 35th day of the epidemic (March 31, 2020), 5,000 on the 42nd day (April 7th) and it will double and reach 10,000 on 46th day of the epidemic (April 11th) whereas until April 11th, the number of cases in the country was just 1825 [37].

## Reliability of predictions on Covid-19 dynamics

### Predictions of cumulative cases

From the selected studies, twenty predicted the cumulative cases for the coming days and 68 predictions were made (Figure 6-a)). The ratio between the predicted value and the observed value was calculated as a criterion for the correctness of the predictions. We found that the predicted values were higher than the observed values for 41.18% of the estimations (28 of the 68) and lower than the observed values for 58.82% remaining estimations (40 of the 68). Only 11 studies (46 estimations) predicted values that actually departed from the values observed in the future (ratio of the predicted value to the observed value less than 0.8 or greater than 1.2). The time periods (expressed as number of days) covered by the data used in the selected studies were considered. From Figure 6-b, the ratio of predicted to actually observed values decreases as the number of days since the first case was reported increases (Linear regression analysis: *β* = –0.124; p=0.002). Thus, the predicted values approach the observed values for lesser time-limited data. There is no difference across the models used to predict the future values of the cumulative cases (Figure 6–c)).

**Figure 6:**
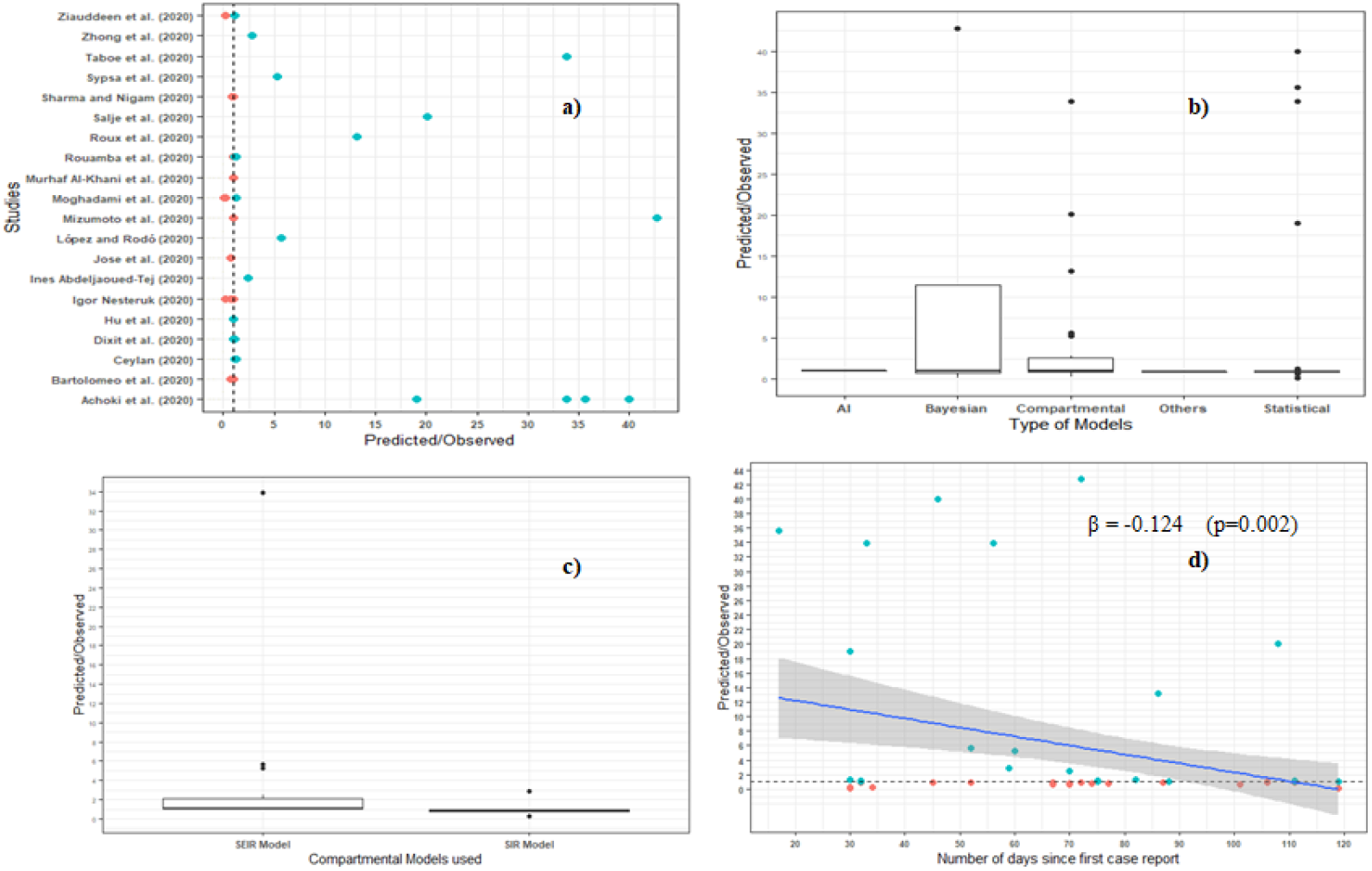
**a)**Ratio of number of cumulative cases predicted to actual number of cumulative cases in 20 studies **b)** Scatter plot of the ratio of number of cumulative cases predicted to actual number of cumulative cases in 6 studies and the number of days covered by the data used for estimation **c**) Boxplot of the ratio of number of cumulative cases predicted to actual number of cumulative cases in 6 studies and the models used

Confidence interval (CI) or credibility interval (CrI) are essential measurements of uncertainties in parameter estimations. As an indicator, the ratio of the amplitude of the 95% CI or 95%CrI and the predicted value was calculated to assess reliability of the estimated values. Very few studies have reported CI or CrI. Only 5.4% of studies (8 out of 148) have reported them in estimating the cumulative number of cases. These 8 studies provided 37 predictions of which, only two had a ratio close to 1 (0.94 and 1.04). The ratio of the amplitude of 95% CI or CrI and the predicted value was greater than 1 for 9 out of the 37 estimations (24.3%) and less than 1 for the 28 remaining (75.7%) (Figure 7-a)). Moreover, this ratio decreases with the length of the period (in number of days) covered by the data used for the estimation (Linear regression analysis: *β* = –0.024; p-value = 0.014, Figure 7-b)). The ratio suggests a difference between the models used (Figure 7-c)). However, this difference cannot be assumed since of the 38 studies that presented a confidence interval, the Bayesian, the compartmental and the statistical models/methods were used in 4, 1 and 31 papers respectively. More data would be needed to better make this comparison.

**Figure 7:**
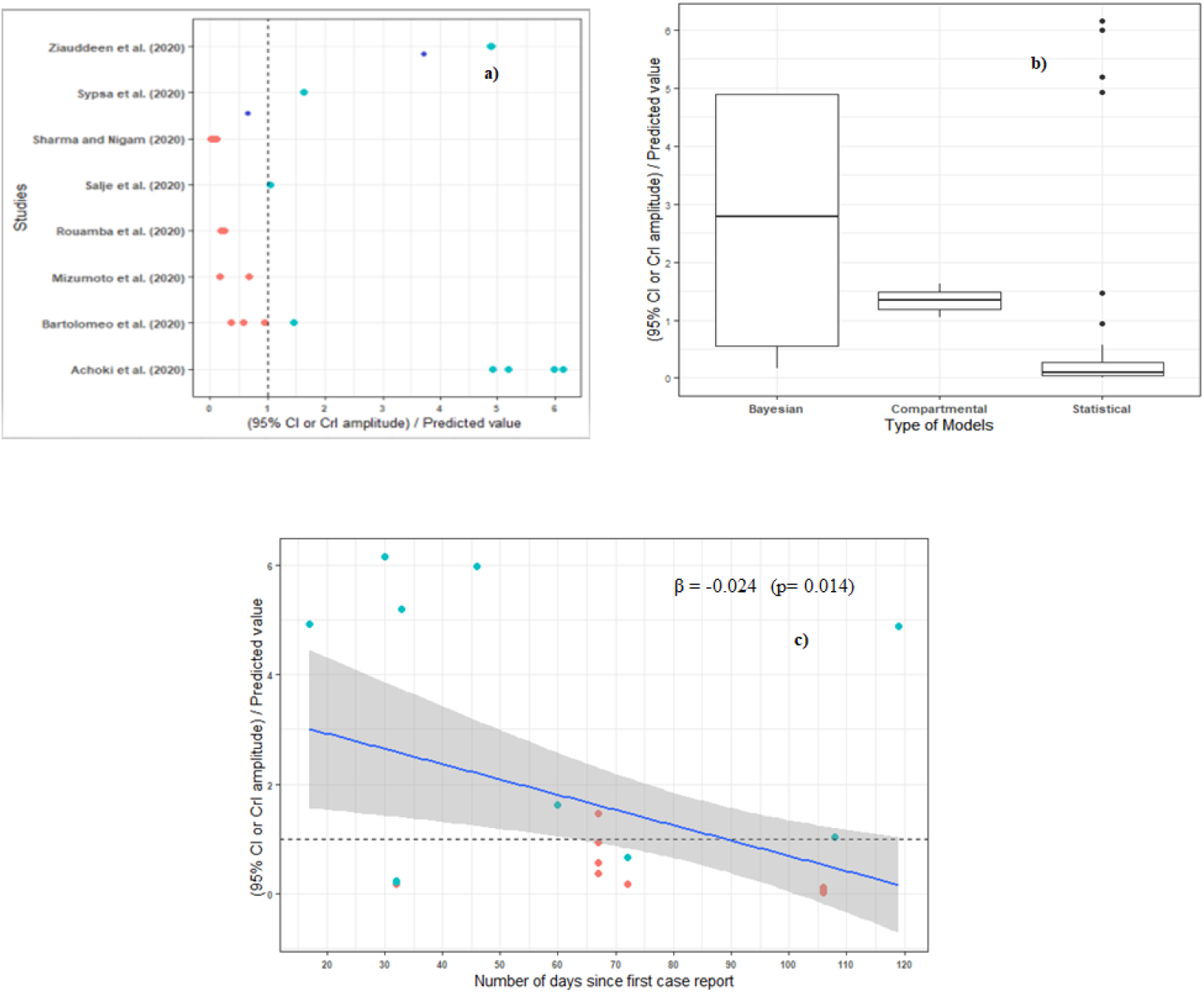
**a)**Ratio of the amplitude of the 95%CI or 95%CrI to the predicted cumulative number of cases in 8 studies **b)** Scatter plot of the ratio of the amplitude of the 95%CI or 95%CrI to the predicted cumulative number of cases and the number of days covered by the data used for estimation **c**) Boxplot of the ratio of the amplitude of the 95%CI or 95%CrI to the predicted cumulative number of cases and the models used

A third indicator was also calculated based on the confidence or credibility intervals provided. This indicator checked whether the true value is within the confidence or credibility intervals provided (Figure 8). Figure 8-a crossed the number of studies that presented a confidence or credibility interval (8 in total) as well as the number of predictions made by each of them with the indicator showing whether or not the true value belongs to the confidence interval. It can be seen from this figure that the majority of the actual values are not found within the confidence or credibility intervals provided (only eight out of 37 of the estimated values were found within them). The true values were found in 6 of the 31 studies that used statistical methods and presented a confidence or credibility interval. The same went for 2 of the 4 studies that used Bayesian methods as well as for the 2 studies that used compartmental models.

**Figure 8:**
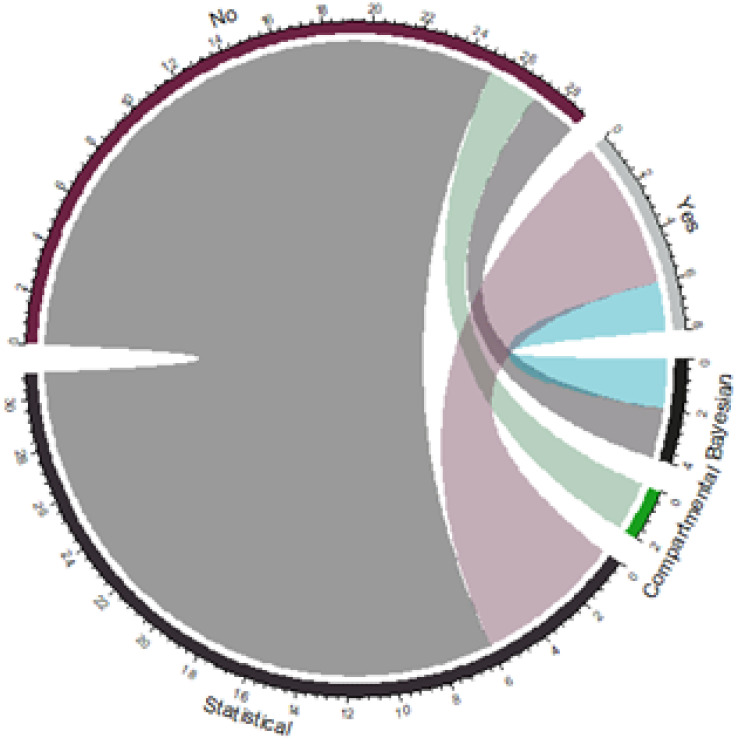
Models used and number of true value of cumulative number of cases that fall within the 95%CI or 95%CrI given in the studies.

## Predictions of deaths

Six of the 148 selected studies (4.05%) predicted the number of deaths with in total 9 predictions. For 33.3% of the predictions, the ratio of predicted to actual number of deaths was close to 1. Two of the nine predictions made a prediction that underestimated the actual number of deaths while four overestimated it (Figure 9-a). Moreover, it appears that the more data used to make the estimates cover a large period of time, the more precise the estimates tend to be, although this is statistically significant only at the 10% threshold (Linear regression Analysis: *β*=–0.030; p-value=0.0945) (Figure 9-b). Figure 9-b meanwhile indicated that the Bayesian model made predictions closer to reality than the compartmental and the statistical models, although these differences are only suggestive due to the small size of the data.

**Figure 9:**
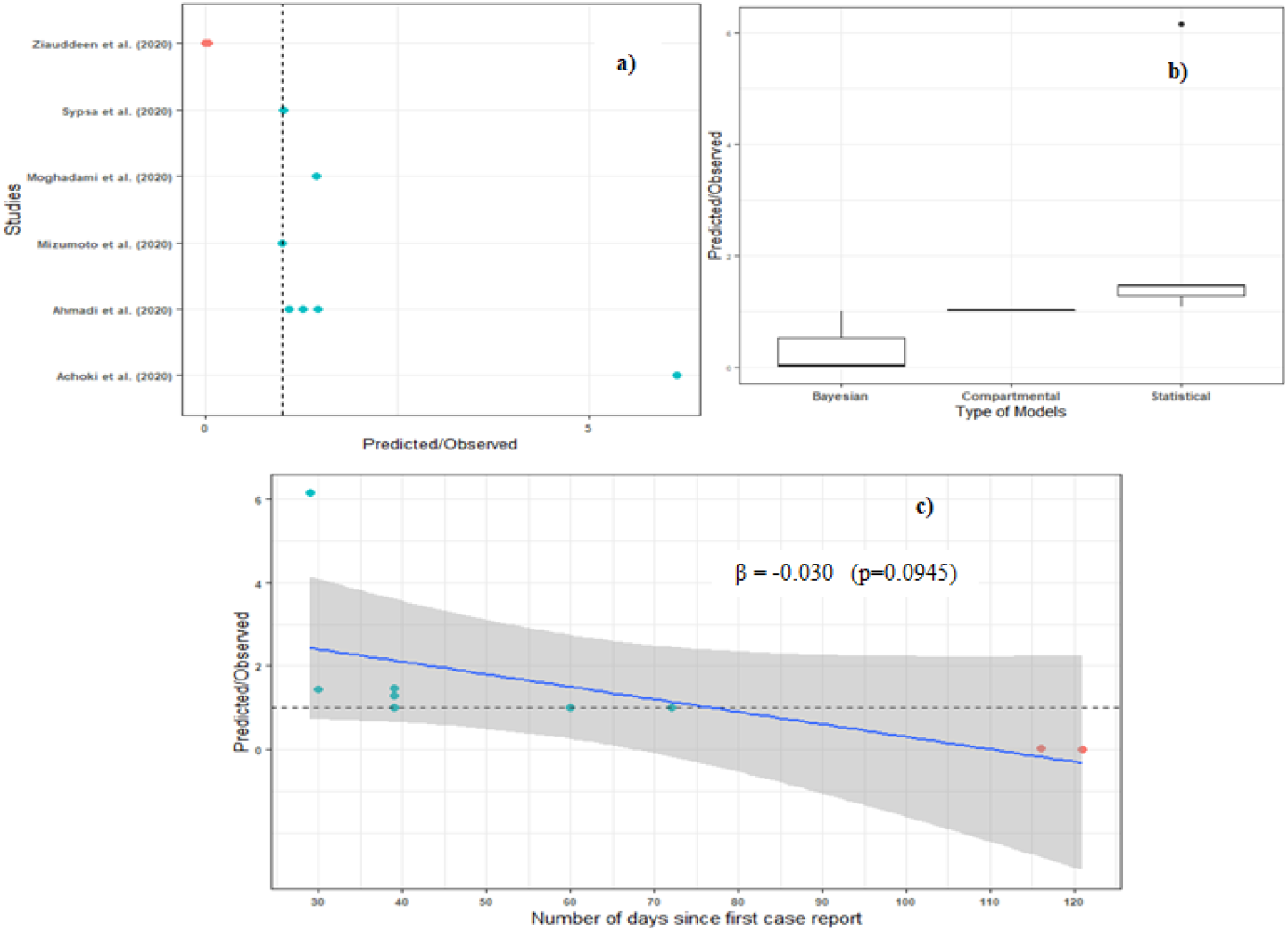
**a)**Ratio of number of deaths predicted to actual number of deaths in 6 studies **b)** Scatter plot of the ratio of number of deaths predicted to actual number of deaths in 6 studies and the number of days covered by the data used for estimation **c**) Boxplot of the ratio of number of deaths predicted to actual number of deaths in 6 studies and the models used.

## Discussion

### Modelling techniques applied to Covid-19 dynamics

The novel coronavirus pandemic (Covid-19) is causing devastating demographic, social, and economic damage globally. Understanding current patterns of the pandemic spread and forecasting its long-term trajectory is essential in guiding policies aimed at curtailing the pandemic [7]. This situation induces a demand to the mathematical epidemiologist community [38] for revealing models of outbreak dynamics, which have not only explanatory but also a predictive potential while the outbreak is in an active phase. These models are aimed at the fast estimations of the future Covid-19 impact on the population, measures required from the public health system and effectiveness of different control measures [39]. A wide-range of models were used; some were extremely simple models while others are more sophisticated. Most studies focused on compartmental models, SIR and SEIR models, to estimate the transmission dynamics and make predictions about the future growth of the pandemic. We found that a SIR model performs better than an SEIR model in representing the information contained in the confirmed-case data though [19] suggested that simpler models may provide less valid forecasts because they cannot capture complex and unobserved human mixing patterns and other time varying characteristics of infectious disease spread. Our finding is in line with that of [17] who reported that predictions using more complex models may not be more reliable compared to using a simpler model. The models used included also statistical models, Bayesian models, Artificial Intelligence based model and Hybrid models [7, 16, 40, 41, 42].

### Reliability of predictions on Covid-19 dynamics

The studies included in this review focused more on the prediction of the cumulative number of cases than the deaths caused by Covid-19. The models used for predicting the dynamics of the Covid-19 were mainly compartmental (SIR and SEIR models) and statistical (Linear regression model, time series model, growth models). We did not find evidence for difference across models while some predicted values far exceed true values. However, our findings should be considered with caution as for the reviewed studies, the number of estimations were not fairly distributed across models. The simplest SIR model is used to predict diseases in which individuals can obtain permanent immunity after infection and is only applicable when there is a non-drug prevention intervention [18]. This model has shown better predictive performance relatively to the SEIR model for number of cases forecasting. Our findings are contrary to that of [18] which argued that the estimated numbers of infected people far exceed reported cases in the available literature which used the SIR model and suggest more the use of complex models. Bayesian models were also used by [16, 43]. Though AI based models were used in few studies of our review, it is shown by [44] that the ongoing development in AI has significantly improved prediction, and forecasting for the Covid-19 pandemic. However, some factors could explain the departure of predictions from actually observed data. Estimates that emerge from modelling studies are only as good as the validity of the epidemiological or statistical models used, the extent and accuracy of the assumptions made and, perhaps most importantly, the quality of the data to which models are calibrated. Early in an epidemic, the quality of data on infections, deaths, tests, and other factors often are limited by under-detection or inconsistent detection of cases, reporting delays, and poor documentation, all of which affect the quality of any model output [19]. In the particular case of Covid-19, [8] argued that the fact that some people only experience mild symptoms and that even the best health system can only detect and treat those presenting to facilities also means that the available data on ‘confirmed’ cases represents only a fraction of the true picture of the pandemic. Additionally, the key to establish a reliable model is to track the epidemic dynamics and release the clinical information and epidemiological data in a timely manner. However, official data are often uncertain because medical resources are limited. The available data only reports confirmed cases in hospitals and ignores infected people who do not have access to medical services. This makes it difficult to accurately predict the course of the epidemic [18, 17]. Moreover, regarding the problem of data quality, especially in developing countries, though we assume that data are collected in real time and adequately in developed countries, this is undoubtedly not the case in underdeveloped countries that do not have the means for doing so. [8], when forecasting cumulative cases, new infections, and mortality due to Covid-19 in Africa found their work further complicated because in Africa, data on key covariates are either lacking or when they exist, they tend to be biased or derived from other global covariate-based modelling exercises. For example, for a growth models at a growth phase of the epidemic, the fact that for 3 - 4 consecutive days no case was reported (often in week-end and 1–2 days after) is not consistent and will certainly introduce bias in predictions.

Another factor that might contribute to the estimations biases is that models are often built on strong assumptions [18] that may not hold. Models may capture aspects of epidemics effectively while neglecting to account for other factors, such as the accuracy of diagnostic tests; whether immunity will wane quickly; if reinfection could occur; or population characteristics, such as age distribution, percentage of older adults with co-morbidities, and risk factors (e.g., smoking, exposure to air pollution) [19]. Additionally, predictions were made in some studies considering scenarios about control measures that do not always match with the reality on the ground. Then the predictions are prone to biases. In most of the studies, parameters estimated from data collected in the first affected countries such as China were used to derive estimates of parameters in other countries [45] even though it is unlikely that epidemics follow identical paths in all regions of the world [19]. An additional point that might also explain the departure of predictions from values actually observed is linked to the fact that predictions are among others intended to guide public health policies for controlling spread of epidemics. As such, based on the predictions, different control measures might have been taken which might have allowed to considerably reduce the number of cases, and hence the observed values, resulting in a “false” overestimation appreciation.

Deterministic models have a long history of application in the study of infectious disease epidemiology. Yet, many infectious disease systems are fundamentally individual-based stochastic processes, and are more naturally described by stochastic models [46]. Therefore, using deterministic models for a stochastic process could also be a source of bias in the estimations. Deterministic model typically describes the average behavior of a system (e.g., populations or sub-populations) without taking into account stochastic processes or chance events in single entities (e.g., individuals). Hence, such models are typically applied to situations with a large number of individuals where stochastic variation becomes less important and heterogeneity can be accounted for using various sub-populations [21]. Stochastic models are models where the parameters, variables, and/or the change in variables can be described by probability distributions. This type of model can account for process variability by taking into account the random nature of variable interactions, or can accommodate parameter uncertainty, and so may predict a distribution of possible health outcomes. Considering process variability can be particularly important when populations are small or certain events are very rare [21]. Stochastic models makes it possible to take into account several factors and lead to a more realistic research. [47] studied the effects of the environment on the spread of Covid-19 using stochastic mathematical model. Modelling studies have contributed vital insights into the Covid-19 pandemic, and will undoubtedly continue to do so [19] although modelling and predicting the epidemiology and trajectory of a disease such as Covid-19 is a challenging exercise

[8]. However, mathematicians and statisticians should be rigorous in their methodology in order to provide robust and reliable results based on which appropriate and optimal management strategies to contain the disease efficiently would be made.

## Conclusion

Using modelling techniques to predict the course of Covid-19 is important to effectively guide public health policy-making. Nevertheless, ensuring that predictions are accurate and correct is necessary to optimize the allocation of the limited resources available, especially in poor countries. Based on the sample of 148 papers analyzed, we showed that compartmental and statistical growth and time series are so far the most used modelling techniques to predict Covid-19 dynamics. Predictions largely exceed the observed values although some predictions (65.21% for cumulative number of cases and 33% for deaths) were close to observed values. We also showed that predictions based on larger dataset (i.e. larger period) were more precise and correct than predictions based on smaller dataset. Ensuring identifiability in model calibrations, data quality, and larger period should provide better confidence in predictions.

## Data Availability

I declare that all data used in the manuscript are available online.

https://ourworldindata.org/coronavirus-source-data

## Acknowledgments

KVS acknowledges the support of the Wallonie-Bruxelles International Post-doctoral Fellowship for Excellence, Belgium (Fellowship N°SUB/2019/443681). RGK acknowledges the support from the African German Network of Excellence in Science (AGNES) and the Alexander von Humboldt Foundation (AvH).

## Appendix

A. Count of selected studies per Countries and Continent

A. Count of selected studies per Countries and Continent
| Continent | Countries | Count | Continent | Countries | Count |
| --- | --- | --- | --- | --- | --- |
| **Africa** | Nigeria | 5 | **Europe** | Italy | 25 |
|  | South Africa | 4 |  | France | 20 |
|  | Algeria | 2 |  | Spain | 15 |
|  | Kenya | 2 |  | Germany | 12 |
|  | Egypt | 1 |  | Uk | 8 |
|  | Senegal | 1 |  | Belgium | 6 |
|  | Burkina-Faso | 1 |  | Netherlands | 5 |
|  | Mauritius | 1 |  | Switzerland | 5 |
|  | Niger | 1 |  | Denmark | 4 |
|  | Tunisia | 1 |  | Sweden | 4 |
|  | Morocco | 1 |  | Slovakia | 3 |
|  | Cameroon | 1 |  | Canada | 3 |
| **America** | US | 15 |  | Norway | 3 |
|  | Brazil | 6 |  | Moldova | 2 |
|  | Peru | 1 |  | Greece | 2 |
|  | Costa-Rica | 1 |  | Bulgaria | 2 |
|  | Mexico | 1 |  | Cyprus | 2 |
|  | Ecuador | 1 |  | Russia | 2 |
| **Asia** | China | 50 |  | Czechia | 2 |
|  | India | 14 |  | Estonia | 2 |
|  | Iran | 5 |  | Finland | 2 |
|  | South Korea | 5 |  | Hungary | 2 |
|  | Japan | 5 |  | Ireland | 2 |
|  | Singapore | 4 |  | Lithuania | 2 |
|  | Korea | 3 |  | Poland | 2 |
|  | Indonesia | 2 |  | Portugal | 2 |
|  | Saudi Arabia | 2 |  | Slovenia | 2 |
|  | Lebanon | 1 |  | Croatia | 1 |
|  | Nepal | 1 |  | Turkey | 1 |
|  | Pakistan | 1 |  | Iceland | 1 |
|  | Malaysia | 1 |  | Latvia | 1 |
| **Oceania** | Australia | 4 |  | Luxembourg | 1 |
|  | Austria | 4 |  | Malta | 1 |
|  | New Zealand | 2 |  | Romania | 1 |

